# Exploring Patient Preference Information in Clinical Research and Decision Models: An Investigative Survey

**DOI:** 10.1101/2024.12.25.24319624

**Authors:** Martina Di Blasio, Zahra Bami, Matteo Bracco, Carmen Fava, Alessia Visconti, Ileana Baldi, Paola Berchialla

**Affiliations:** Center for Biostatistics, Epidemiology, and Public Health, Department of Clinical and Biological Sciences, University of Turin, Italy; Department of Clinical and Biological Sciences, University of Turin, Italy; Department of Cardiac Thoracic Vascular Sciences and Public Health, University of Padua, Italy

**Keywords:** Patient preference information, Survey, Evidence synthesis, Decision-making

## Abstract

Patient preferences are playing an increasingly pivotal role in defining care pathways, assessing care quality, and advancing healthcare technologies, but their current use remains unclear. We conducted an online survey collecting data from 46 experts in clinical research, pharmaceutical industry, regulatory affairs, and health technology assessment. About 30% of our respondents are using PPI routinely, and 90% are willing to prioritize their integration in the future, despite various challenges, such as limited patient engagement, difficulty in obtaining reliable data, and reluctance to deviating from traditional practices. We show here a growing recognition of the potential benefits of integrating PPI, and that concerted efforts and investments are necessary to support a wider adoption.

## Introduction

Patients are increasingly recognized as key stakeholders in healthcare decision-making^1–3^. Hence, the integration of patient preferences information (PPI) in becoming critical^4,5^, particularly when multiple therapeutic options lack clear distinctions in efficacy. In such scenarios, the patient’s perspective on side effect profiles, treatment administration methods, and/or impact on daily life can be the deciding factor in treatment selection^6^. This ensures that medical decisions align with clinical best practices and consider individual patients’ values and lifestyle, likely enhancing treatment adherence, health outcomes, and patient satisfaction^7^. This is in line with the concept of value-based healthcare, which focuses on delivering the care that matters most to patients while ensuring efficient resource utilization^8^.

To ensure medical products are developed and evaluated to effectively meet patient needs from conception to market and beyond, and to inform regulatory decisions to ensure approved treatments align with patient needs and desires^9–11^, PPI must be considered in decision-making throughout the entire medical product life cycle, from early research and development to clinical trials, regulatory decisions, and post-market surveillance^12–14^. However, the extent to which healthcare professionals understand and apply knowledge from patient preference studies remains unclear. To investigate the integration of PPI into clinical research and new treatments development as well as to explore the awareness, understanding, and interests related to PPI, we present here the results of a comprehensive online survey targeting experts in clinical research, pharmaceutical industry, regulatory affairs, and health technology assessment, providing the readers with suggestions for improving PPI use in healthcare decision-making.

## Material and methods

### The survey

The survey, available as **Supplementary Data 1**, was designed to investigate how patient preferences are perceived, utilized and valued. To this aim, we first conducted a literature search to identify the main domains in which preference studies are applied, *i*.*e*., clinical trial design, treatment decision-making, and drug development. Then, we used these domains to directly inform the wording of our survey items.

The survey was administered via Typeform, and was open from April 1st, 2023, to June 30th, 2023. It included 16 questions divided in six key areas: *Frequency of Use*: respondents were asked to indicate how often they incorporate patient preference information in their work, ranging from “ never” to “ always”; *Perceived Benefits*: the survey explored the perceived advantages of integrating patient preferences, such as improved trial design, enhanced patient recruitment and retention, and more patient-centric outcomes; *Implementation Challenges*: participants were prompted to identify an8d rate the significance of various obstacles to integrating patient preferences, including methodological complexities, resource constraints, and regulatory uncertainties; *Organizational Readiness*: questions assessed the availability and accessibility of patient preference data within respondents’ organizations, as well as the existence of processes for incorporating this information; *Current Practices*: the survey inquired about specific ways in which patient preference information is currently being used in respondents’ work, such as in protocol design, endpoint selection, or benefit-risk assessments; and *Future Outlook*: respondents were asked about their expectations for the future role of patient preferences in clinical research and treatment development.

### Survey respondents

We used a snowball sampling approach, starting from respondents from SIMeF fellows (https://www.simef.it/). They were then invited to forward the invitation to any colleagues who might be interested in taking part, and those colleagues were asked to do the same, resulting in a total of 47 respondents. To encourage the response rate^15^, respondents were remunerated with €10, which they could either receive as an Amazon voucher (n = 24) or donate to the Red Cross (n = 21). Two respondents decided not to receive or donate any remuneration.

No personal information (*e*.*g*., age, sex, geographical location) was collected. We included one question to verify that users were paying attention to the questionnaire (*i*.*e*., “ We want to verify that you are paying attention to the questions in the questionnaire, so we ask you to select the option “ three”) which disqualified one respondent.

### Statistical analysis

Descriptive statistics (*i*.*e*., absolute and relative frequencies) were generated using base R (v 4.4.2). Figures were generated using *ggplot2* (v 2.3.5, bar plots in Figure 1 and 2), *ggsankey* (v 0.0.99999, Sankey plot on Figure 1), and *ggsuburst* (v 0.3.0, Sunburst plot in Supplementary Figure 1).

**Figure 1.**
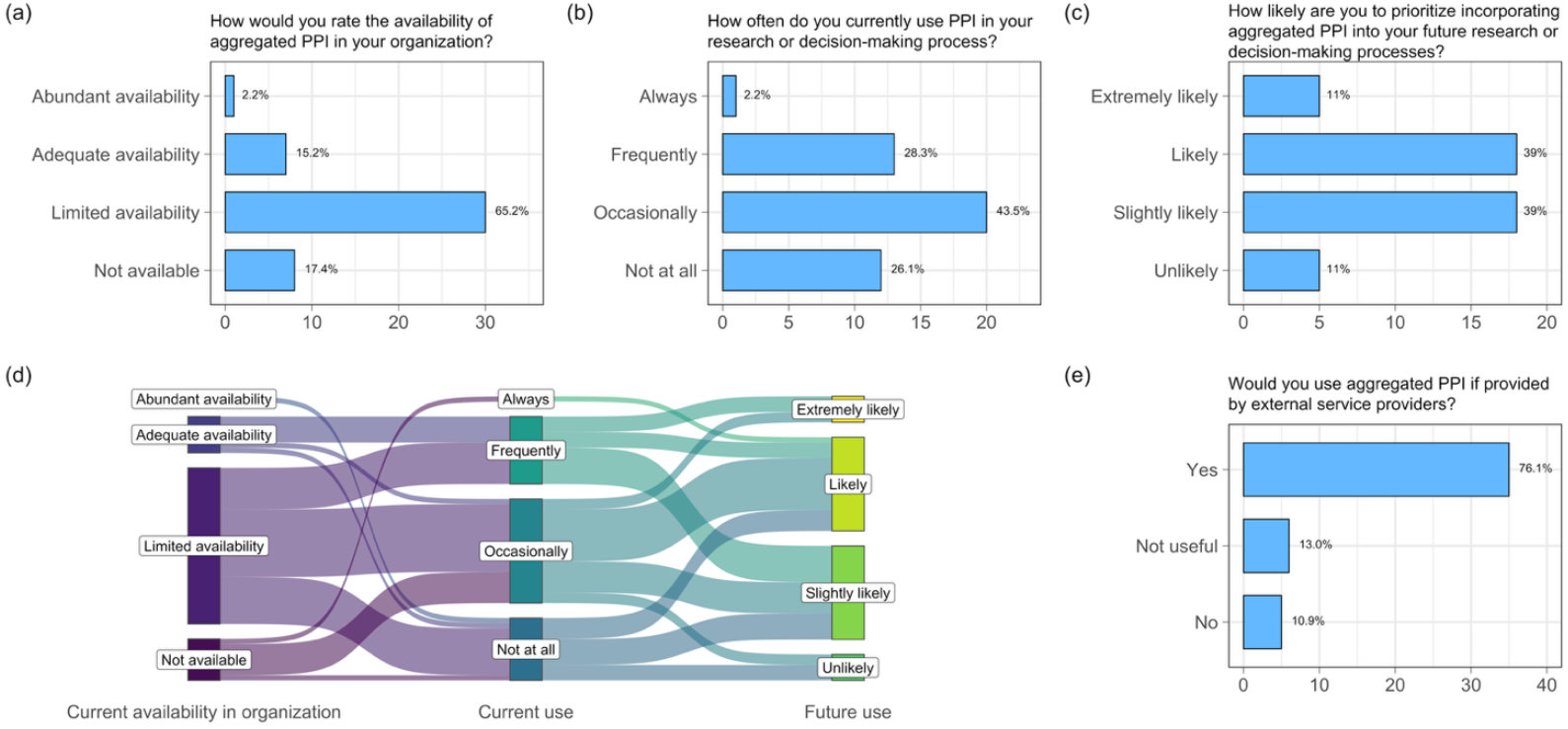
Patient Preference Information: Use in Research and Development. The bar plots in the upper panel show the distribution of responses regarding the (a) current availability and (b) usage of PPI as well as (c) the likelihood of a future usage, with the (d) Sankey plot showing the flow of answers between these three topics. The bar plot on the bottom panel (e) shows the distribution of responses regarding the desire for external service providers supplying PPI. In the bar plots, the x-axes show absolute frequencies, with relative frequencies being reported beside each bar. All questions allowed a single answer. PPI: patient preference information.

**Figure 2.**
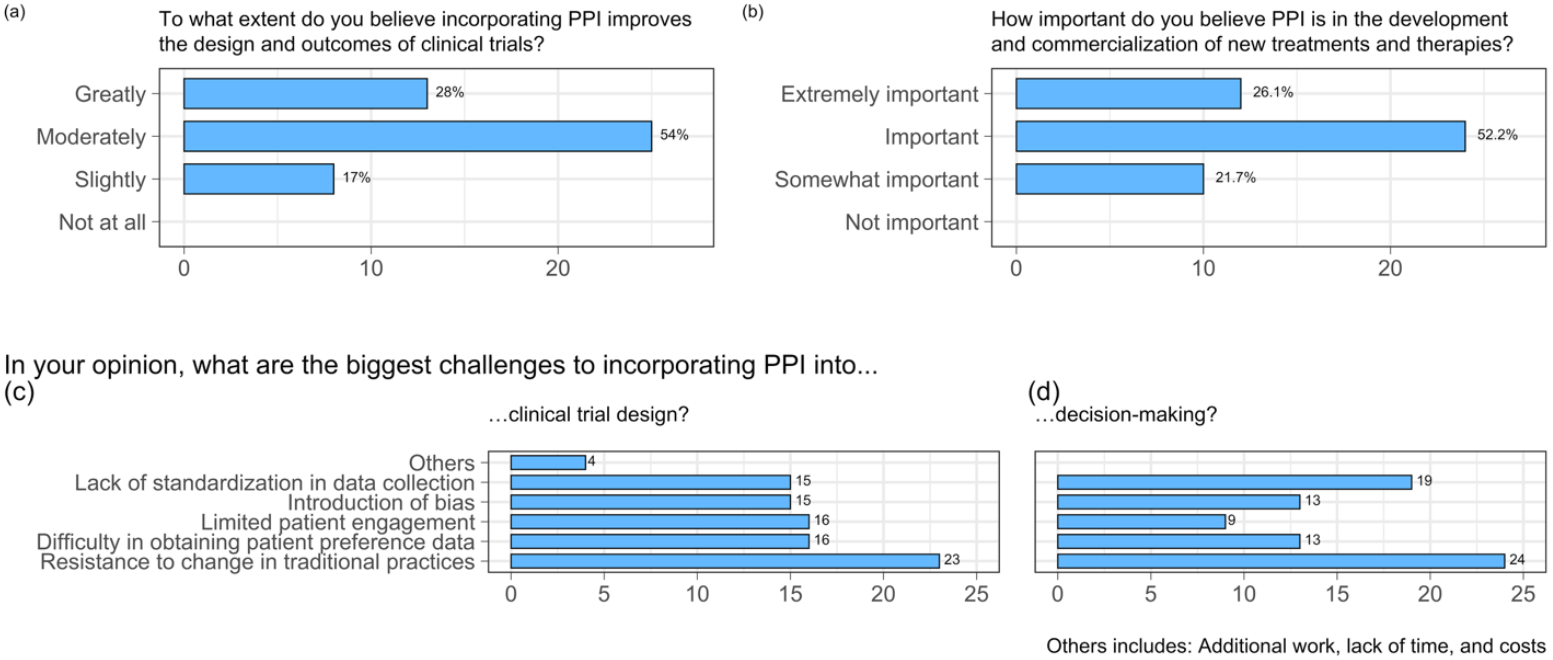
Patient Preference Information: Importance and Challenges in Research and Development. The bar plots show the distribution of responses regarding the importance of PPI in (a) the design and outcomes of clinical trials and (b) development and commercialization of new treatments and therapies as well as the challenges that researchers predict facing by including them in (c) clinical trials design and (d) decision-making. The x-axes show absolute frequencies. The questions presented in the upper panels allowed a single answer, and their relative frequencies are reported beside each bar. The questions presented in the bottom panel allowed multiple answers, and their total absolute frequencies are reported beside each bar. PPI: patient preference information.

Due to the explorative nature of this study, the sample size was not pre-determined, and all the received questionnaires were analysed, with no inclusion or exclusion criteria. Answers were processed anonymously in aggregate, and in compliance with GDPR regulations.

## Results

### Participant Characteristics

We elicited valid responses from 46 Italian participants hailing from various sectors, including pharmaceutical and biotechnology companies (n = 27, 58.7%), academic and research institutions (n = 6, 13.0%), hospitals and clinics (n = 5, 10.9%), government agencies (n = 2, 4.3%), consulting firms (n = 1, 2.2%) and vendors (n = 1, 2.2%), with respondents also at the intersection between multiple roles (i.*e*., academic and research and hospitals and clinics, n = 4, 8.7%). The primary occupational domains encompassed researchers (n = 10, 21.7%), healthcare professionals (n = 9, 19.6%), statisticians (n = 8, 17.4%), quality assurance (n = 3, 6.5%), policy or decision analysts (n = 2, 4.3%), pharmacovigilance experts (n = 2, 4.3%), as well as various management and consultancy roles (n = 11, 23.9%), and a patient advocate (2.2%).

About half of the respondents researched on more than one disease (n = 21, 45.7%), with slightly less than one quarter of them researching on more than three areas (n = 11, 23.9%). Neurological, cardiovascular and oncological diseases were the most active research areas, with at least 10 respondents mentioning them, followed by endocrine, respiratory, gastrointestinal and renal diseases, infectious diseases and vaccines, and chronic pain (n respondents ≥ 5, **Supplementary Figure 1**).

### Patient preference information: current and future use

The availability of PPI within organizations is at best limited, with only 8 respondents reporting it to be at least adequate (17.5%; **Figure 1a**). In line with this observation, most respondents reported using PPI only occasionally or not at all (n = 32, 69.6%; **Figure 1b**), though future usage is likely (n = 41, 89.1%; **Figure 1c**). Notably, the lack of availability of PPI within one’s organization is not a clear indicator of its actual use: 7 out of the 8 respondents whose organizations did not provide any patient preference information still use it at least occasionally (**Figure 1d**), confirming the inclination of using such data when provided by external services (n = 35, 76.1%; **Figure 1e**). We also noted that the current use of PPI in research or decision-making processes positively influences future use (**Figure 1d**), highlighting the significance of such data.

### Patient preference information: importance and challenges

Respondents agree that PPI could improve the identification of innovative clinical trials endpoints, at least moderately (n = 38, 82.0%, **Figure 2a)**, and play a crucial role for the development and commercialization of new treatments and therapies (n = 36, 73.8%, **Figure 2b**), and would always have an impact, even if limited. However, significant ideological and practical challenges remain in effectively incorporating these preferences into practice, mostly due to the resistance to innovation, and limited patient engagement, with the consequent difficulty in obtaining preference data (**Figure 2c** and **2d**). Interestingly, lack of time and/or budget and the necessity of additional work represent only a small challenge for the use of PPI in the clinical trials design (**Figure 2c)**, but not in the decision-making process (**Figure 2d**).

## Discussion

Understanding and incorporating patients’ values, priorities, and trade-offs in healthcare decision-making, thus recognizing that truly effective personalized care goes beyond clinical outcomes, has become a crucial aspect of developing and evaluating medical products^5,7,9^. However, data on the integration of PPI in clinical research and in the development of new treatments is lacking.

Here, by using data collected across various stakeholder groups, we observed that, while half of respondents expressed an inclination to prioritize the integration of PPI in their future research projects, the overall current utilization rates are suboptimal, with 26% of respondents reporting that they never employ PPI, and an additional 44% doing so only occasionally. This is partially due to the inadequate availability of aggregated patient preference, with 65% of respondents reporting limited availability, and 17% indicated a complete lack of such data. Therefore, we advocate here for concerted efforts and investments to establish reliable data repositories, as 76% of participants expressed interest in using them, if provided.

Further challenges hinder effectively incorporating PPI into healthcare delivery and product development, as already observed by previous literature^16–18^. Respondents mentioned the lack of standardized data collection methodologies as one of the main problems. This highlight the need to enhance awareness regarding past and current efforts in data collection and standardization, such as the Patient-Centered Outcomes Research Institute’s (PCORI) methodology standards^19^, and the IMI-PREFER project^5^. IMI-PREFER, which was mentioned only by one of our respondents, developed a robust framework that outlines the methodology for conducting patient preference studies^20^, including guidelines for study design^21,22^, and stakeholder interaction^11^. Two of its key aspects are the standardization of the methods used to collect and analyse PPI, including defining qualitative (*e*.*g*., discrete choice experiments, best-worst scaling, and conjoint analysis), and quantitative (*e*.*g*., in-depth interviews, focus groups, and Delphi method) techniques, and creating templates and guidelines for conducting studies in diverse disease areas^19,20^.

Respondents further reported limited patient engagement, difficulty in obtaining reliable preference data, and concerns regarding the potential introduction of bias as important challenges. This builds on stereotypes, with, for instance, many professionals perceiving patient preferences as subjective or less relevant compared to clinical data and scientific evidence^17^. We believe that creating direct communication channels (*e*.*g*, round-table discussions, transparent reporting, feedback loops) between pharmaceutical companies and disease-specific patient advocacy groups and/or global patient networks, which already have strong connections and trust within the patient community, will challenge these stereotypes, and foster a more collaborative environment. Additionally, we believe that providing patients with educational materials about the relevance of their input in healthcare decision-making can further strengthen their willingness to participate actively in shaping research and regulatory priorities.

More worrisomely, about half of the respondents cited as major obstacle the resistance to deviating from traditional practices. Shifting mindset is not trivial, and much effort will be needed. For instance, it has already been recognized that traditional evaluation methods prioritize quantitative data and economic metrics, especially when resources are limited, and financial considerations may overshadow the need for a patient-centered approach^16^. We advocate here for regulatory bodies, such as the US Food and Drug Administration and the European Medicines Agency, to encourage industry stakeholders to adopt PPI systematically. For instance, they could allow expedited pathways, particularly in cases where patient preferences highlight unmet medical needs or shape clinically meaningful endpoints. Additionally, we believe that companies should establish dedicated roles, thus ensuring that patient preference data are appropriately collected, analysed, and applied to decision-making. An increasing exposure to best practices will contribute to a cultural shift, encouraging a more receptive and informed approach.

Despite these challenges, about 90% of the respondents expressed at least a mild inclination to prioritize integrating patient preference information in their future research or decision-making processes. This positive outlook reflects a growing recognition of the potential benefits of adopting a more patient-centric approach in future research or decision-making processes.

While our survey was grounded in the existing literature on patient preference studies, it is important to note that our findings may have underlying bias due to the relatively small sample size. However, such number of respondents is not uncommon, with other similar studies on multiple stakeholders involving between 12 and 143 individuals^12,18,23^. Further research involving larger and more diverse populations is necessary to validate and generalize our findings.

## Conclusion

Our study provides a much-needed overview of the current state of utilizing patient preferences in clinical research and decision-making processes.

We observed that, while the adoption of PPI remains limited, there is a growing recognition of its potential benefits and increasing interest in integrating it into future processes, despite existing challenges and disparities in the utilization rates across organizations and stakeholder groups. Concerted efforts are however needed to overcome identified challenges, such as resistance to change, limited patient participation, lack of standardization and the availability of high-quality data. A patient-centered approach is essential for promoting a more effective, equitable and patient-focused healthcare system.

## Data Availability

Data collected during the study are available to bona fide researchers under managed access due to governance constraints, and can be requested to the corresponding author.

## Data sharing statement

Data collected during the study are available to *bona fide* researchers under managed access due to governance constraints, and can be requested to the corresponding author.

## Ethical Review Board Statement

We confirm that the procedures comply with national and EU legislation. Research was performed in accordance with the Declaration of Helsinki. Ethics committee of the University of Turin (Comitato di Bioetica d’Ateneo) waived ethical approval as well as the need of obtaining informed consent for this work (Protocol number: 0722874) since we used data which were anonymizes before the analysis was performed, and did not collect any sensitive information. By returning a completed questionnaire, respondents consented to be included in the study.

## Acknowledgements

This work was partially supported by “ NODES Spoke 5 project” no ECS00000036, which has received funding from MUR-M4C2 1.5 of PNRR (code BERP_PNRR_EI_23_01_F). This work was partially supported by the “ Proof of Value Instrument 2022 Initiative” (S1921_POV_D218) funded by Compagnia San Paolo and University of Turin.

## Disclosure

The authors declare no competing interests.

## Author Contributions

MdB, IB, and PB conceptualized and designed the project. MdB, CF, IB, and PB designed the survey. MdB, AV, and PB carried out the statistical analyses with contributions from ZB. AV prepared the figures. MdB, AV, IB, and PB interpreted the results, with contributions from MB. MdB, AV, and PB wrote the manuscript. All authors read and approved the final manuscript.

## Notes

### Competing Interest Statement

The authors have declared no competing interest.

### Summary of Updates

We edited the test for clarity, shortening the Introduction and adding details to the Methodology. We also improved the Discussions. No new analyses were performed.

