## Supplementary Material for "Exploring Patient Preference Information in Clinical Research and Decision Models: An Investigative Survey"

### Supplementary Figures

In which disease area is your research focused?

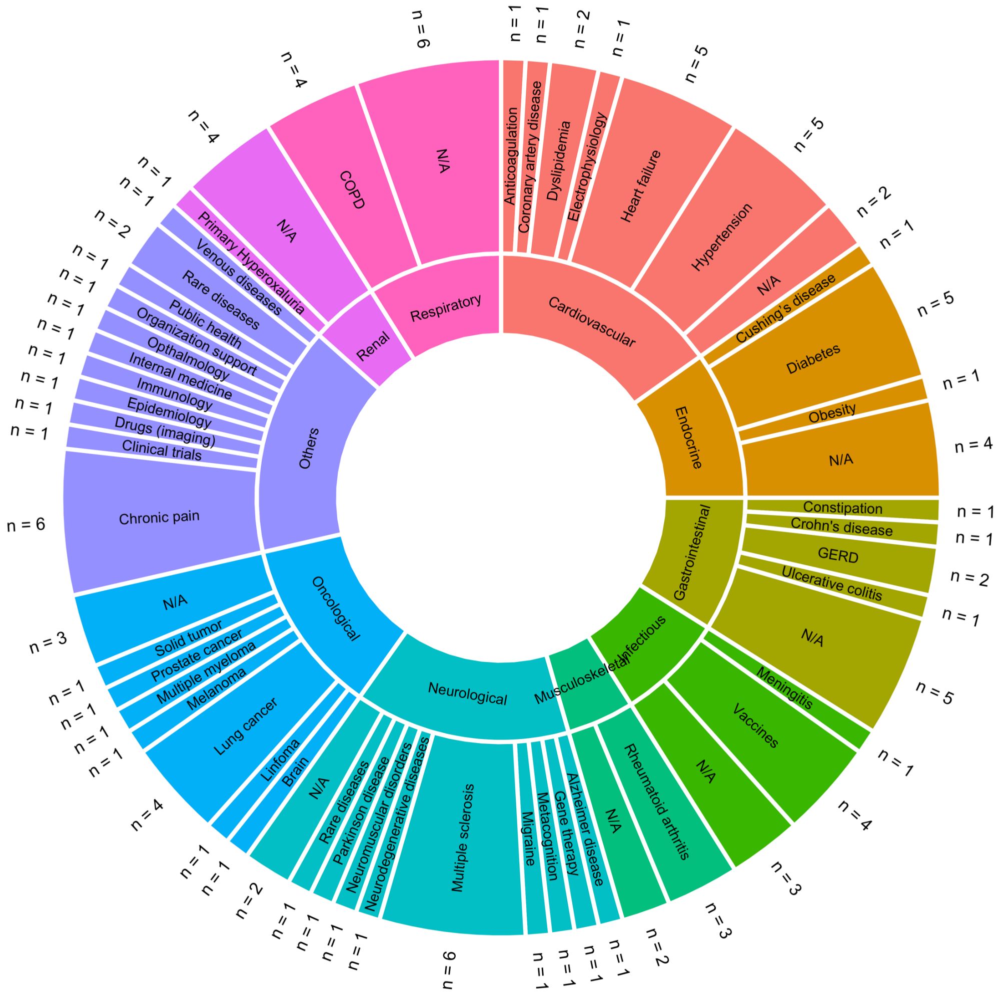


Abbrevoations: GERD: Gastroesophageal reflux disease, COPD:Chronic obstructive pulmonary disease

**Supplementary Figures 1.** The sunburst plot shows the main (inner circle) and specific (outer circle) disease areas in which respondents' research is focused. Slices are coloured according to the main research area. When a respondent did not specify any specific disease, this is counted as N/A. Multiple answers were allowed, and their total absolute frequencies are reported.

### Supplementary Data

**Supplementary Data 1.** Questions included in the questionnaire.

1. In which setting do you primarily work?
   a. Academic/research institution
   b. Hospital/clinic
   c. Pharmaceutical/biotech company
   d. Government agency
   e. Other (please specify)

2. What is your current job title or role?
   a. Researcher
   b. Clinician
   c. Healthcare professional
   d. Policy/decision analyst
   e. Other (please specify)

3. In which disease area is your research focused?
   a. Cardiovascular diseases
      i. Heart failure
      ii. Other
   b. Endocrine diseases
      i. Diabetes
      ii. Other
   c. Gastrointestinal diseases
   d. Mental health disorders
   e. Musculoskeletal diseases
      i. Rheumatoid arthritis
      ii. Other
   f. Neurological diseases
      i. Neuromuscular disorders
         1. Lateral amyotrophic sclerosis
         2. Multiple sclerosis
         3. Other
      ii. Other
   g. Oncological diseases
      i. Lung cancer
      ii. Multiple myeloma
      iii. Other
   h. Renal diseases
   i. Respiratory diseases
      i. Chronic Obstructive Pulmonary disease (COPD)
      ii. Other
   j. Other
      i. Chronic pain
      ii. Gene therapy in haemophilia
      iii. Other, please specify:

4. How often do you currently use patient preference information in your research or decision-making process?
   a. Not at all
   b. Occasionally
   c. Frequently
   d. Always

5. To what extent do you believe incorporating patient preference information improves the design and outcomes of clinical trials?
   a. Not at all
   b. Slightly
   c. Moderately
   d. Greatly

6. How important do you believe patient preference information is in the development and commercialization of new treatments and therapies?
   a. Not important
   b. Somewhat important
   c. Important
   d. Extremely important

7. In your opinion, what are the biggest challenges to incorporating patient preference information into clinical trial design?
   a. Lack of standardization in data collection
   b. Limited patient engagement
   c. Difficulty in obtaining patient preference data
   d. Resistance to change in traditional practices
   e. Introduction of bias
   f. Other (please specify)

8. In your opinion, what are the biggest challenges to incorporating patient preference information into decision-making?
   a. Lack of standardization in data collection
   b. Limited patient engagement
   c. Difficulty in obtaining patient preference data
   d. Resistance to change in traditional practices
   e. Introduction of bias
   f. Other (please specify)

9. How would you rate the availability of aggregated patient preference data in your organization?
   a. Not available
   b. Limited availability
   c. Adequate availability
   d. Abundant availability

10. How likely are you to prioritize incorporating aggregated patient preference information into your future research or decision-making processes?
   a. Unlikely
   b. Slightly likely
   c. Likely
   d. Extremely likely

11. In your opinion, how aggregated patients preference data could be used by academic/research institutions?

12. In your opinion, how aggregated patients preference data could be used by HTA agencies?

13. In your opinion, how aggregated patients preference data could be used by pharmaceutical/biotech companies?

14. In your opinion, how aggregated patients preference data could be used by government agencies?

15. Would you use aggregated data if provided by external service providers?
   a. Yes
   b. No
   c. Not useful

16. Do you have any additional comments or suggestions regarding the use of patient preference information in research and decision-making?
